# Does choice of athletic footwear affect musculoskeletal injury risk in US Coast Guard recruits? A retrospective cohort study

**DOI:** 10.1101/2022.01.03.22268706

**Authors:** John J. Fraser, Ryan Pommier, Andrew J MacGregor, Amy Silder, Todd C. Sander

## Abstract

**Context:** Musculoskeletal injuries (MSKIs) are ubiquitous during initial entry military training, with overuse injuries in the lower extremities the most frequent. A common mechanism for overuse injuries is running, an activity that is an integral part of United States Coast Guard (USCG) training and a requirement for graduation.

**Objective:** Assess the effects of athletic footwear choice on lower quarter MSKI risk in USCG recruits.

**Design:** Descriptive Epidemiological Study

**Setting:** USCG Training Center, Cape May, NJ

**Participants:** A retrospective cohort study was performed in which 1229 recruits (1038 males, 191 females) were allowed to self-select athletic footwear during training. A group of 2876 recruits (2260 males, 616 females) who trained under a policy that required obligatory wear of prescribed athletic shoes served as a control.

**Main Outcome Measures:** Demographic data and physical performance were derived from administrative records. Injury data were abstracted from a medical tracking database. Multivariable logistic regression was used to assess group, age, sex, height, body mass, and run times on MSKI outcomes.

**Results:** Ankle-foot, leg, knee and lumbopelvic-hip complex diagnoses were ubiquitous in both groups (experimental: 20.37 to 29.34 per 1000 recruits; control: 18.08 to 25.59 per 1000 recruits). Group was not a significant factor for any of the injuries assessed. Sex was a significant factor in all injury types, with female recruits demonstrating ∼2.00 greater odds of experiencing running-related injuries (RRIs), overuse injuries, or any MSKI in general. When considering ankle-foot or bone stress injuries, the risk in female recruits was 3.73 to 4.11 greater odds than their male counterparts. Run time was a significant predictor in RRI, all overuse injuries, and for any MSKI in general.

**Conclusion:** While footwear choice did not influence MSKI risk in USCG recruits, female sex was a primary, nonmodifiable intrinsic risk factor.

**Key Points:**

- Self-selection of athletic footwear was not a significant factor for MSKI in USCG recruits, despite widespread reports of discomfort and perceived deleterious effects of wear.
- MSKI continues to be a major source of morbidity in the recruit training population, with female sex found to be a robust and significant factor regardless of footwear choice.
- While there is no indication that prescribed athletic footwear is associated with MSKI, policymakers should factor in individual preference when considering the mandate for wear.

## INTRODUCTION

Musculoskeletal injuries (MSKIs) are ubiquitous during initial entry military training, impacting 12–22% of all recruits,^1–3^ with overuse injuries in the lower extremities the most frequent.^4–6^A common mechanism for overuse injuries is running,^7^ an activity that is an integral part of United States Coast Guard (USCG) training and a requirement for graduation. Therefore, it is important to understand the factors, including the impact of footwear, that may contribute to MSKI risk. Athletic footwear forms the interface between the foot and ground during running and is hypothesized to affect running-related injuries (RRIs) through control of ankle-foot kinetics and kinematics.^8^ More recently, the focus of injury risk has been on modifiable movement patterns such as foot strike type, cadence and running surface, all of which can affect factors related to injury risk, such as average and peak loading rate, peak hip adduction, and knee flexion angle at landing.^9–12^ While footwear type (i.e., minimalist, motion control, etc.) often affects running mechanics,^10,13^ a direct association between running footwear and injury risk has yet to be established.^8,14,15^ It has been suggested that self-selection of comfortable shoes with a preferred movement path is more closely associated with injury prevention.^8,16^

In 1941, the US Congress passed the Berry Amendment to promote domestic commerce through the obligated purchase of American-made products by the military, which includes uniforms. Historically, athletic footwear was not considered a uniform item. However, the 2017 National Defense Authorization Act included an interpretation that Berry Amendment-compliant athletic footwear be included with the enlisted recruit uniform issue to “provide sufficient choices to minimize the incidence of athletic injuries in initial entry training.”^17^ In response to this determination, the USCG Training Center (TRACEN) began mandating the use of prescribed athletic footwear in the first quarter of 2018 and required all recruits to wear them during training, regardless of their preferred footwear or prior running experience. Additionally, scheduling density during the recruit training pipeline typically precluded the necessary time to adequately assess fit and comfort during uniform issue or acclimatize recruits to the footwear prior to training, which could plausibly contribute to increased MSKI risk.^8,18,19^

Acknowledging that self-selection of athletic footwear may influence MSKI in the recruit population while still ensuring adequate footwear was provided to recruits regardless of their pre-enlistment socioeconomic status, a proof-of-concept project was initiated in October 2019 that allowed recruits a choice to either wear the prescribed Berry-compliant athletic footwear or their own footwear. Due to the potential for this policy change to influence injury outcomes, empirical assessment of the effects of this policy is warranted. Therefore, the purpose of this retrospective cohort study was to assess the effects of athletic footwear choice on lower quarter MSKI risk, while factoring the influences of age, sex, height, body mass, and physical fitness level as assessed by run time from the Physical Fitness Assessment (PFA). A secondary aim was to assess the perceived form, fit, and function of prescribed uniform footwear in all recruits.

## METHODS

This was a retrospective cohort study that included a sample of recruits stationed at the USCG TRACEN, Cape May, New Jersey, from March 2018 to March 2020. Ethics approval was provided by the USCG Institutional Review Board as exempt research. The Strengthening the Reporting of Observational studies in Epidemiology (STROBE)^20^ was used to guide reporting of this study.

### Data Sources

Recruit age, sex, height, body mass, and run time on the PFA were derived from administrative records maintained by USCG TRACEN. Injury data, including the diagnosis, body segment, and the attributed mechanism of injury, were abstracted from a medical tracking database maintained by USCG TRACEN medical personnel. Additionally, data from a process improvement project, which solicited feedback from all recruits on the form, fit, and function of the issued athletic footwear, were also examined.

### Cohort Selection

A sample of 2876 recruits (2260 males, 616 females) stationed at USCG TRACEN from 01 October 2018 to 30 September 2019, who trained under the policy that required obligatory wear of Berry-compliant athletic shoes, were included in the control group. **Figure 1** details the prescribed uniform running footwear manufactured by New Balance Athletics, Inc. (Boston, MA, USA) and San Antonio Shoemakers (SAS; San Antonio, TX, USA) for the US Department of Defense that were available at USCG TRACEN during the study epoch. A sample of 1229 recruits (1038 males, 191 females) stationed at USCG TRACEN from 01 October 2019 to 31 March 2020, a time where a shift in administrative policy allowed for the use of self-selected athletic footwear during training, comprised the experimental group. The exposure was consistent for both cohorts as the training curriculum, mode, and frequency of physical training at USCG TRACEN were similar during the study epoch. Since several intrinsic risk factors may contribute to lower quarter injury,^21^ age, sex, height, body mass, and PFA run scores were collected.

**Figure 1.**
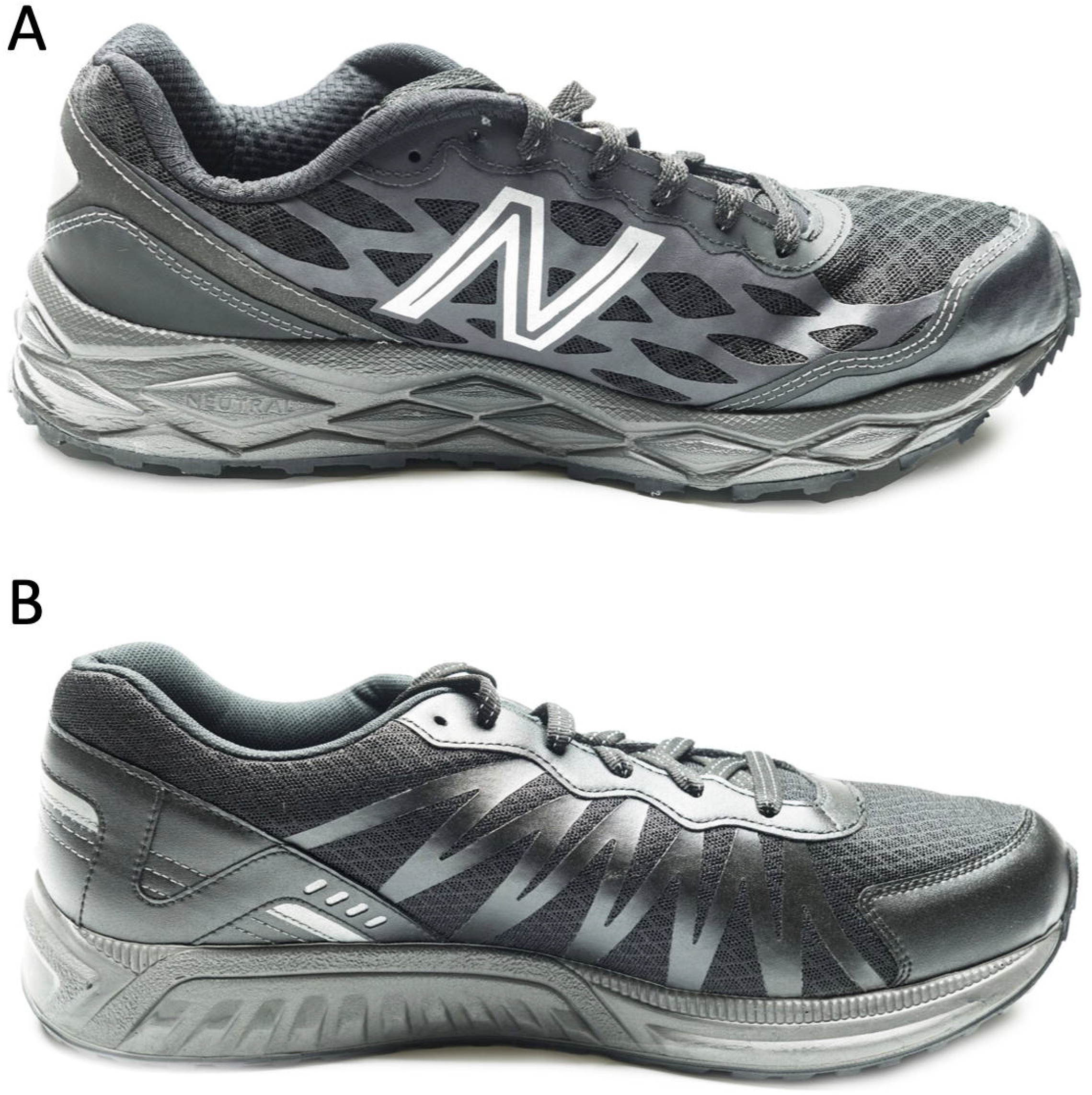
Prescribed uniform running footwear manufactured by (A) New Balance Athletics, Inc (Boston, MA) and (B) San Antonio Shoemakers (SAS; San Antonio, TX)

### Outcome Variables

Recruits who presented to the medical treatment facility at TRACEN for any lower quarter MSKI during training were counted as having an outcome. Since footwear choice may influence certain types of injuries over others, characteristics of the injury type (e.g., bone stress injury and medial tibial stress syndrome [MTSS]), body region (e.g., hip, knee, ankle-foot), and attributed mechanism (e.g., running) were derived from the medical database. For this study, the specific diagnostic categories of interest included ankle-foot complex injuries (specifically involving the tibia, fibula, ankle, or foot), MTSS or bone stress injuries (involving any bone in the lower quarter), RRI (any lower quarter injury specifically attributed to running), overuse injuries (any lower quarter injuries that were not caused by a macrotraumatic event), and all MSKIs (which encompassed both repetitive microtraumatic and macrotraumatic injuries). Given that these categories are not mutually exclusive, a recruit with an injury could potentially be counted in one or multiple categories. For example, a recruit that presented for a tibial bone stress injury from running would be categorized in each of the five categorizations of injury outcomes employed in this study. In contrast, a recruit with anterior knee pain experienced during squatting would only be classified in the overuse injuries and all MSKI categories. If no clear mechanism was identified by the recruit, idiopathic MSKI was assumed to be a result of a repetitive overuse injury.^22^

### Perceived Form, Fit, and Function of Berry-Compliant Prescribed Athletic Footwear

A process improvement project was conducted from April 2019 to September 2019 that solicited feedback pertaining to the form, fit, and function of the prescribed uniform athletic footwear among all USCG recruits (n=922) upon graduation. These recruits were asked to rate the level of satisfaction with the issued athletic footwear (5-point Likert scale: 1=very dissatisfied, 3=neutral, 5=very satisfied); if they ever suffered any pain with these shoes, (yes/no); have they ever suffered an injury as a result of wearing these shoes (yes/no); how much of an effect did the shoes have on their running performance (5-point Likert scale: 1=ran significantly slower, 3=no effect, 5=ran significantly faster); and which of the following footwear would you prefer (issued uniform athletic footwear or self-selected habitual athletic footwear).

### Statistical Approach

The level of significance was *p* < 0.05 for all analyses. Descriptive statistics were calculated for both the experimental and control groups. To evaluate the effects of the prescribed athletic footwear on MSKI risk, a multivariable logistic regression was performed modeling the effects of experimental group (referenced to prescribed footwear controls), age, female sex (referenced to male recruits), height, body mass, PFA run scores, and the interactions of experimental group by body mass and experimental group by female sex. Analyses were performed using the survival package (version 3.2-10) in R version 3.5.1 (The R Foundation for Statistical Computing, Vienna, Austria). Odds ratios (ORs) and 95% confidence intervals (CIs) were calculated for each significant predictor using the exponentiated log odds. For the secondary aim of assessing the results of feedback pertaining to the form, fit, and function of the prescribed footwear, descriptive statistics of responses were reported.

## RESULTS

**Figure 2** details the group demographics. The control group had greater mean age (≤1 year) and a lower proportion of females that comprised the group (control group: 15.7%; experimental group: 21.4%). There were no other statistically significant differences in group demographics. When stratified by sex, there were significant differences in recruit characteristics, with female recruits exhibiting greater mean age (≤1 year), shorter height, lower body mass, and longer run times.

**Figure 2.**
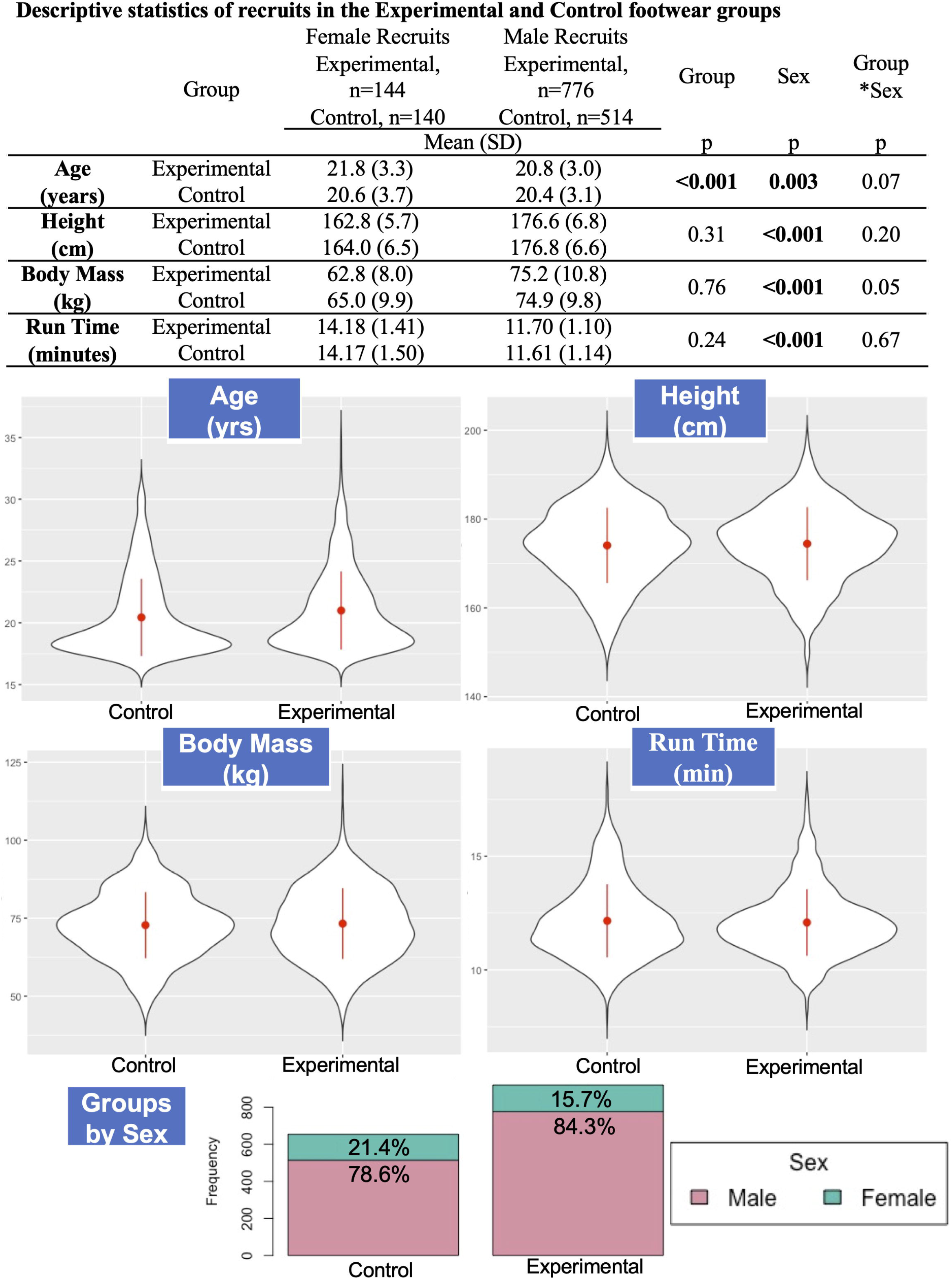
Descriptive statistics of recruits in the Experimental and Control footwear groups

**Figure 3** details the prevalence of MSKI in the experimental and control groups. There were no significant differences in MSKI between groups for any of the injuries or injury mechanisms reported (**Table 1**). Ankle-foot, leg, knee, and lumbopelvic-hip complex complaints were ubiquitous in both groups, ranging from 20.37 to 29.34 per 1000 recruits in the experimental group and 18.08 to 25.59 per 1000 recruits in the control group. Thigh injuries were comparatively scarcer, with a prevalence of 4.89 per 1000 recruits in the experimental group and 2.73 per 1000 recruits in the control group. When characterizing the type of injuries, overuse injuries were predominant (experimental group: 96.17 per 1000 recruits; control group: 86.32 per 1000 recruits) compared with traumatic injuries (experimental group: 11.41 per 1000 recruits; control group: 5.46 per 1000 recruits). Among overuse injuries, RRIs were a substantial burden in both the experimental (72.53 per 1000 recruits) and control (64.14 per 1000 recruits) groups. MTSS and bone stress injuries occurred in about one third of RRIs. When considering all injuries, the burden was 107.58 per 1000 recruits in the experimental group and 91.78 per 1000 recruits in the control group.

**Table 1.**
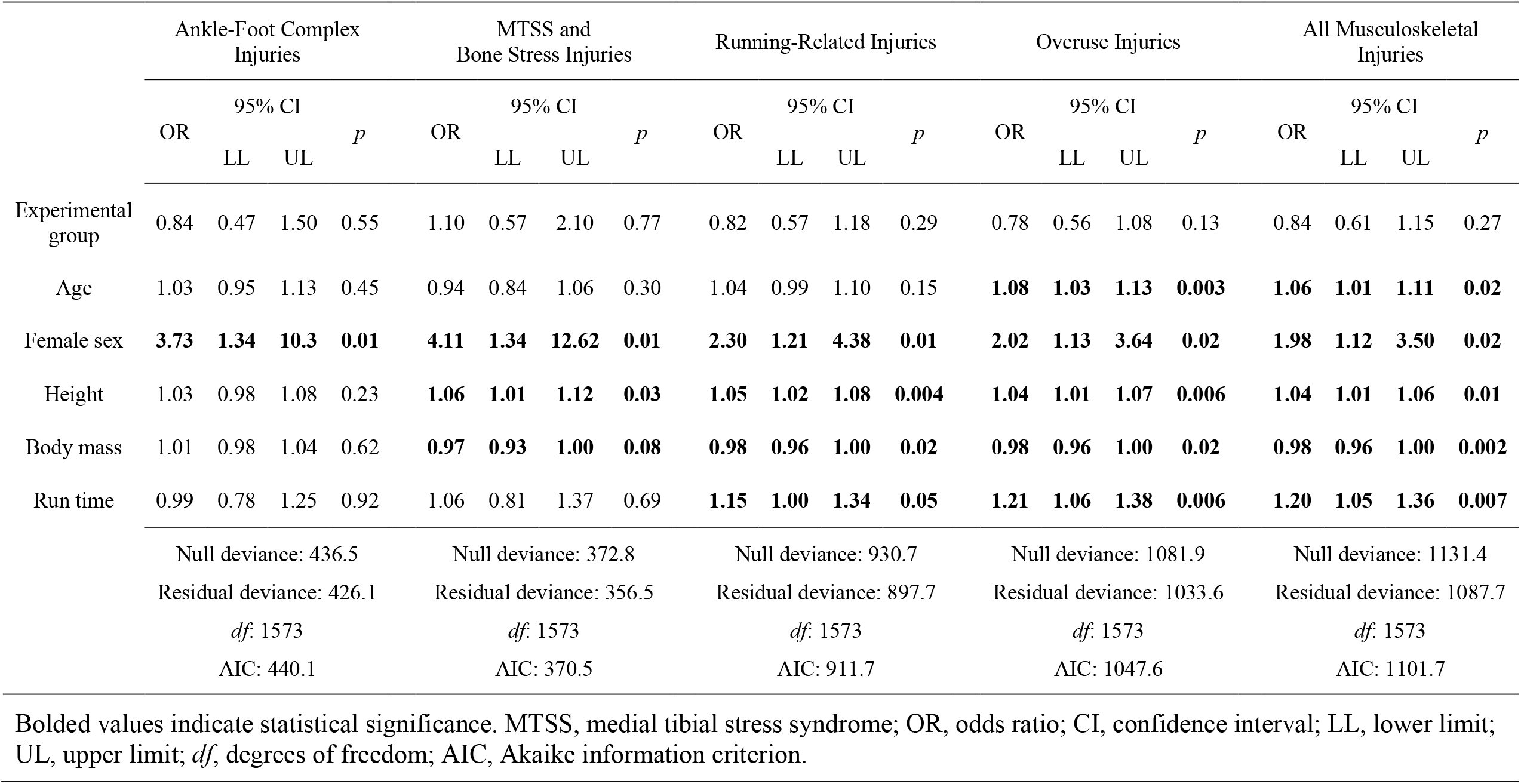
Results of the multiple variable logistic regression analyses modeling musculoskeletal injuries in US Coast Guard recruits

**Figure 3.**
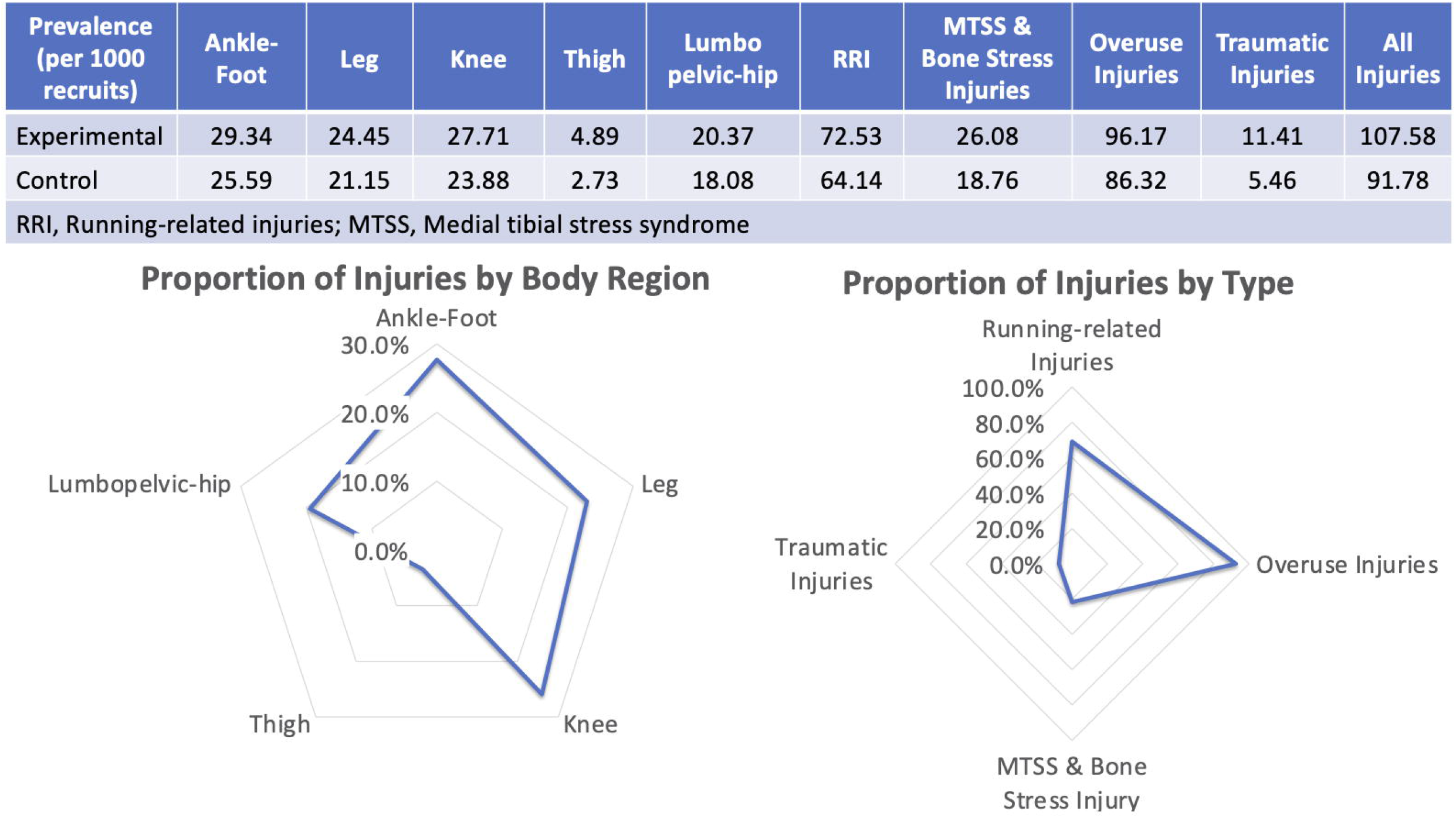
Prevalence of MSKI in the experimental and control groups.

The results of the multivariable logistic regression analysis are reported in **Table 1**. Group was not a significant factor for any of the injuries assessed. Sex was a significant factor in all injury types, with female recruits demonstrating ∼2.00 greater odds of experiencing an RRI, overuse injuries, or any MSKI in general. When considering ankle-foot, MTSS, or bone stress injuries, the risk in female recruits increased to 3.73 to 4.11 greater odds than their male counterparts. Run time was a significant predictor in RRI, all overuse injuries, and for any injury (OR range 1.15–1.21. A significant but small magnitude increased risk was observed for the factors of greater age and a small magnitude protective effect was observed for greater body mass. The interactions of experimental group by body mass and experimental group by female sex were not significant (OR and CI crossed a 1.00 threshold, *p*>0.05) for any injury type and were removed from the final models.

**Table 2** details the results of the survey pertaining to the form, fit, and function of prescribed footwear in all recruits. The majority (58.6%) of the respondents reported pain and 15.7% attributed an MSKI to wearing the prescribed uniform footwear. Pertaining to perceived effect on running performance, approximately one quarter of the respondents reported a deleterious effect, 10.4% reported an advantageous effect, and 63.3% reported no effect. Approximately one quarter of the respondents reported satisfaction with the prescribed footwear, approximately one half reported being dissatisfied, with the remaining one quarter reporting neutrality. The prescribed uniform footwear was the preferred footwear of 29.3% of the respondents, with only 12.3% reporting continued use of the footwear following graduation.

**Table 2.** Survey of form, fit, and function of prescribed uniform footwear for all recruits

|  |  | New Balance |  | San Antonio Shoewear |  | Total |  |
| --- | --- | --- | --- | --- | --- | --- | --- |
|  |  | (n=869) |  | (n=53) |  | (n=922) |  |
|  |  | n | % | n | % | n | % |
| Reported pain during wear |  | 506 | 58.2 | 34 | 64.2 | 540 | 58.6 |
| Injuries attributed to the prescribed footwear | Ankle-foot complex | 120 | 13.8 | 10 | 18.9 | 130 | 14.1 |
|  | Knee | 16 | 1.8 | 1 | 1.9 | 17 | 1.8 |
|  | Thigh | 2 | 0.2 | 0 | 0.0 | 2 | 0.2 |
|  | Lumbopelvic-hip complex | 10 | 1.2 | 1 | 1.9 | 11 | 1.2 |
|  | Total | 135 | 15.5 | 10 | 18.9 | 145 | 15.7 |
| Perceived effect on running performance | Substantially slower | 42 | 4.8 | 4 | 7.5 | 46 | 5.0 |
|  | Slower | 176 | 20.3 | 11 | 20.8 | 187 | 20.3 |
|  | No effect | 550 | 63.3 | 34 | 64.2 | 584 | 63.3 |
|  | Faster | 85 | 9.8 | 4 | 7.5 | 89 | 9.7 |
|  | Substantially faster | 16 | 1.8 | 0 | 0.0 | 16 | 1.7 |
| Satisfaction with prescribed uniform footwear | Very satisfied | 51 | 5.9 | 6 | 11.3 | 57 | 6.2 |
|  | Somewhat satisfied | 197 | 22.7 | 12 | 22.6 | 209 | 22.7 |
|  | Neutral | 214 | 24.6 | 10 | 18.9 | 224 | 24.3 |
|  | Somewhat dissatisfied | 205 | 23.6 | 11 | 20.8 | 216 | 23.4 |
|  | Very dissatisfied | 197 | 22.7 | 14 | 26.4 | 211 | 22.9 |
| Preferred footwear | Prescribed | 256 | 29.5 | 14 | 26.4 | 270 | 29.3 |
|  | Control | 613 | 70.5 | 39 | 73.6 | 652 | 70.7 |
| Intended continued use of prescribed footwear following graduation |  | 108 | 12.4 | 5 | 9.4 | 113 | 12.3 |

## DISCUSSION

This study retrospectively investigated whether MSKI risk differed between USCG recruits who were uniformly prescribed Berry-compliant athletic footwear versus those who were free to choose their own athletic footwear. We found that choice of athletic footwear had no significant effect on MSKI in USCG recruits. This is despite the widespread report of discomfort and dissatisfaction of the prescribed athletic footwear observed in the feedback provided by the recruits. Our study also confirms that MSKI continues to be a major source of morbidity in the recruit training population, with female sex found to be a robust and significant factor regardless of footwear choice.

Use of the prescribed athletic footwear did not increase the risk of MSKI, despite reported perceived attribution. These findings are consistent with high-level evidence that the use of prescribed footwear in the recruit and active duty military populations is neither protective nor contributory to MSKI.^18,23^ This also supports the findings of Bullock and colleagues^24^ who concluded that that there was insufficient evidence to recommend prescribed footwear given that arch height and replacement of footwear at regular intervals did not significantly influence MSKI. Saragiotto and colleagues^25^ studied perceptions of the role of foot morphology and athletic footwear in recreational runners and found that about one third of the participants attributed foot morphology to RRI and viewed footwear as especially salient for prevention.^25^ Some of these same beliefs were expressed by Wolthon and colleagues^26^ who reported continued belief in the contribution of footwear toward MSKI among both physical therapists and nonclinicians. Despite the dissatisfaction with the prescribed athletic footwear in our current study and individuals’ perceived association of footwear and MSKI reported by others studies,^25,26^ our results support the conclusions of many others^8,14,15,18,23,24^ that athletic footwear appears to have no significant influence on MSKI. It is highly plausible that the perceptions reported by the USCG recruits may be shaped through social mechanisms, especially if other recruits were vocal about their dislike of the footwear. This may explain the disparity between the widespread subjective discomfort and lack of objective association of the prescribed footwear with MSKI observed in our study. This supposition, and some of the qualitative factors that contribute to dissatisfaction, warrant further investigation.

Our findings align with a previous report on a similar training population that identified female sex as a significant factor for MSKI. Cosio and collegaues^27^ conducted a study of USCG cadets during Summer Warfare Annual Basic Training and found increased odds in female recruits for ankle (OR: 2.58), foot (OR: 2.07), bone stress (OR: 1.13), and overuse (OR: 1.72) injuries. Slower run times were also found to be associated with MSKI in the same study.^27^ Our findings also diverge from a recent systematic review with meta-analysis that found while sex was a factor for MSKI in female military members, it was not a significant factor for repetitive overuse injuries.^21^ We observed large and significant effects for female sex in each injury type in the multivariable assessment, including specifically MTSS and bone stress injury (OR: 4.11, 95% CI: 1.34–12.62), RRI (OR: 2.30, 95% CI: 1.21–4.38), and overuse injuries (OR: 2.02, 95% CI: 1.13–3.64). These findings also agree with an increased risk of ankle-foot complex fractures observed among Navy enlisted females compared with their male counterparts.^28^

### Clinical and Policy Recommendations

While there is no indication that the Berry-compliant prescribed athletic footwear is associated with MSKI, policymakers should factor in individual preference when considering the mandate for wear. Based on the reported dissatisfaction, perceived discomfort, and the low proportion of Coast Guardsmen who intended to continue use of the prescribed footwear beyond the 8-week recruit training, return on investment for this appropriation should be considered. To balance the domestic commercial benefits of purchasing Berry-compliant prescribed athletic footwear with the individual needs of the service member, it is recommended that all stakeholders (recruit training leadership, instructor cadre, and recent recruits) be involved during the contracting process by wear-testing and providing feedback to establish best value. This will help ensure continued use of issued athletic footwear beyond recruit training and maximize the return on investment of appropriated funds.

While the schedule at recruit training is constrained and precludes time for the recommended sufficient transition period to the new footwear,^29^ allocating time for proper fitting, distribution, and acclimatization to the footwear upon arrival at TRACEN could remedy this barrier. Lastly, it is also recommended that the instructor cadre and clinicians at TRACEN clearly communicate to recruits that there is no reliable evidence linking athletic footwear with the prevention (or contribution to development) of RRI.^29^ This may help to dispel any preconceived beliefs or attitudes toward the prescribed footwear.

There are limitations to this study. Consistent with USCG records management policies, administrative records are routinely destroyed at the end of the life cycle. As such, a portion of the administrative records containing the demographic information of height, mass, and PFA run times of both groups were not available at the initiation of this study. While this precluded analysis of all recruits during this study epoch and may have introduced bias, these records were destroyed randomly and a large representative sample was nevertheless included.

## CONCLUSION

Self-selection of athletic footwear was not a significant factor for MSKI in USCG recruits, despite widespread reports of discomfort and perceived deleterious effects of wear. MSKI continues to be a major source of morbidity in the recruit training population, with female sex found to be a robust and significant factor to these outcomes regardless of footwear choice.

## Data Availability

Due to the nature of this research, participants of this study did not agree for their data to be shared publicly, so supporting data is not available.

